# A new perspective on the BO06 trial in osteosarcoma: short- and long-term prognostic value of histologic response and intensified chemotherapy

**DOI:** 10.1101/2021.05.14.21255716

**Authors:** Eni Musta, Nan van Geloven, Jakob Anninga, Hans Gelderblom, Marta Fiocco

## Abstract

**Purpose:** Cure rate models accounting for cured and uncured patients, provide additional insights into long and short term survival. We aim to better understand the prognostic value of histologic response and chemotherapy intensification on cure fraction and progression-free survival (PFS) for the uncured patients.

**Methods:** A logistic model is assumed for the effect of histologic response and intensified chemotherapy on the cure status, while a Cox regression model is estimated only for the uncured patients on PFS. The mixture cure model is used to simultaneously study these two effects.

**Results:** Histologic response is a strong prognostic factor for the cure status (OR: 3.00 [1.75-5.17]), but it has no clear effect on PFS for the uncured patients (HR: 0.78 [0.53-1.16]). The cure fractions are 55% [46%-63%] and 29% [22%-35%] among patients with good histologic response (GR) and poor responders (PR) respectively. The intensified regimen was associated with higher cure fraction among PR (OR: 1.90 [0.93 – 3.89]), with no evidence of effect for GR (OR: 0.78 [0.38 – 1.59]).

**Conclusions:** Accounting for cured patients is valuable in distinguishing the covariate effects on cure and PFS. Estimating cure chances based on these prognostic factors is relevant for counseling patients and can affect treatment decisions.

## Background

Osteosarcoma is a malignant bone tumor that occurs mostly in children, adolescents and young adults. Surgery alone is insufficient for curation of osteosarcoma patients and the introduction of adjuvant chemotherapy has led to significant improvements in survival.^1-2^ Current treatment for osteosarcoma includes neoadjuvant chemotherapy in combination with adequate surgery and different treatment schedules have been used in the past to establish an optimal chemotherapeutical design.^3-5^ Recently, in a randomized controlled international collaborative study, the European and American Osteosarcoma Study Group (EURAMOS) concluded that the 3 drug-regimen, consisting of Methotrexate, Adriamycin (Doxorubicin) and Cisplatin (MAP) is the standard treatment. In the experimental arms no survival benefit of introducing Interferon-α-2b, an immune-modulator drug, for good responders (GR, < 10% viable tumor cells after induction chemotherapy),^6^ or intensifying treatment by adding Ifosfamide and Etoposide for poor responders (PR,≧ 10% viable tumor cells after induction chemotherapy)^7^ could be established. With respect to dose intensity, defined as given dose in mg/m^2^ per time unit^8^, a phase-III study (BO06) in osteosarcoma that randomized patients between a conventional dose-intensity (CI) regimen or dose-intensified (DI) regimen with the same drug combination (Doxorubicin plus Cisplatin) in both arms, showed no difference in outcome^9^. Despite the fact that the group of patients with the DI-regimen had a higher proportion of GR, no better event free or overall survival was seen in this group^9^. It is noteworthy that histologic response, as assessed by histologic examination of the surgical specimen, is used as a key prognostic factor for survival of patients with osteosarcoma. Among PR, 5-year PFS is around 50% and among GR it is over 70%.^7,10^ However, histologic response has been a debate as a surrogate marker for outcome in osteosarcoma.^6,9-11^ So, if even this actor is considered to be important, its interpretation is difficult, particularly in relationship to dose-intensity, and it is of interest to understand how this, and other factors contribute to better survival of PR.

In osteosarcoma, there is an ongoing debate about the effect of doses and dose intensities,^4,9,12-14^ which is an issue for other cancer types as well.^15-22^ All these studies have identified the presence of long-term disease-free survivors among osteosarcoma patients, who are practically considered as ‘cured’. Late relapse after 5 years follow-up occurs only in less than 5% of the patients.^23,24^ This suggests, at the moment that primary treatment is completed, a patient actually belongs to one of two categories: the cured ones and the uncured, who will experience disease progression within their lifetime. However, the identification of cured patients can only be done after a patient has been observed to remain disease-free for many years after treatment. In particular for patients alive and progression-free who have only been followed for a few years, the category to which they belong is not known. Hence all the patients are usually studied together as one group. However, in the presence of a significant fraction of cured patients, the effect of a treatment or other prognostic factors may relate either to the probability of never experiencing progression or to the time free from progression for the uncured. This distinction is not captured by the traditional methods, i.e. it is not possible to identify whether survival has improved since the treatment is curing more patients or because the uncured are living longer. As a result, certain effects might not be identified.

For this reason, cure rate models have started to be adopted in oncological studies as an alternative statistical modelling approach, which by accounting for cured and uncured patients, can provide additional insights into long and short term survival patterns.^25-31^ Through these models, it is possible to identify prognostic factors with a cure or a life-prolonging effect.^25,32^ In addition to survival probabilities as in the traditional framework, cure models also allow for estimation of a cure fraction, the chance that a given patient is cured, and of the PFS for the uncured patient. Such information may be used to select patients at high risk of progression for more aggressive chemotherapeutical strategies and protect those with high chances of being cured from the toxic side effects.

In this study, the BO06 clinical trial^9^ is revisited from a new point of view focusing on two different survival outcomes: the cure fraction and the PFS for the uncured patients. By accounting for cured patients, the prognostic value of histologic response and intensification of chemotherapy is evaluated on each of these outcomes with the aim to reveal new insights into the complex nature of such effects.

## Methods

### Patients and chemotherapy information

Data from the MRC BO06/EORTC 80931 randomized controlled trial for patients with localized resectable high-grade osteosarcoma, diagnosed between May 1993 and September 2002 were considered.^9^ Patients were randomly assigned to the conventional two-drug regimen (Reg-C), consisting of six 3-week cycles of doxorubicin (DOX, 75 mg/m2) and cisplatin (CDDP, 100 mg/m2), or to the dose-intense regimen (Reg-DI), consisting of same doses administered 2-weekly and supported by G-CSF (5 μg/kg daily). The two-week cycles of Reg-DI correspond to increasing the planned dose intensity of DOX and CDDP by a factor 1.5 compared to the conventional 3-week cycles. For both groups, surgery was scheduled at week 6 since the start of treatment, i.e. after 2 cycles for Reg-C and after 3 cycles for Reg-DI, and postoperative chemotherapy was intended to resume 3 weeks after surgery. However, the received course of chemotherapy was often different from the intended one as a result of delays and reductions related to treatment-induced toxicity. The dataset consists of 497 patients (245 assigned to Reg-C and 252 to Reg-DI) aged 40 years or less. More details about the patients and chemotherapy can be found in the primary publication of the trial^9^.

### Sample selection and follow-up

The outcome of interest is progression-free survival (PFS), i.e. time free from any relapse (including distant metastasis) or death. PFS is computed from end of treatment because cure, if it occurs, may happen at any time during treatment and assuming that patients are either cured or not at early stages post-diagnosis it would not be appropriate. In addition, covariates such as histologic response and received dose or dose-intensity are not known yet at time of randomization. End of treatment is computed from the starting date of the last chemo cycle received, by adding the planned duration of the last cycle (14 or 21 days depending on the treatment arm). If the first follow-up visit after treatment occurs before this date, the first follow-up visit is considered as end of treatment (meaning that the treatment was interrupted or completed earlier than planned). For patients who do not receive any additional chemotherapy after surgery the date of surgery is considered as the end of treatment. Patients who did not receive chemotherapy, reported an abnormal dosage of one or both agents (more than 1.25×prescribed dose),^13^ did not have surgery or histologic response was not reported, died or had disease progression during the treatment period, were excluded from the original dataset. The consort diagram is given in Figure 1. A total of 379 patients were included in the analysis. Since the focus of the study is investigating cure or cancer progression after primary treatment, the overall survival is not further considered in this study because it incorporates also the effects of further treatments after cancer relapse.

**Fig. 1.**
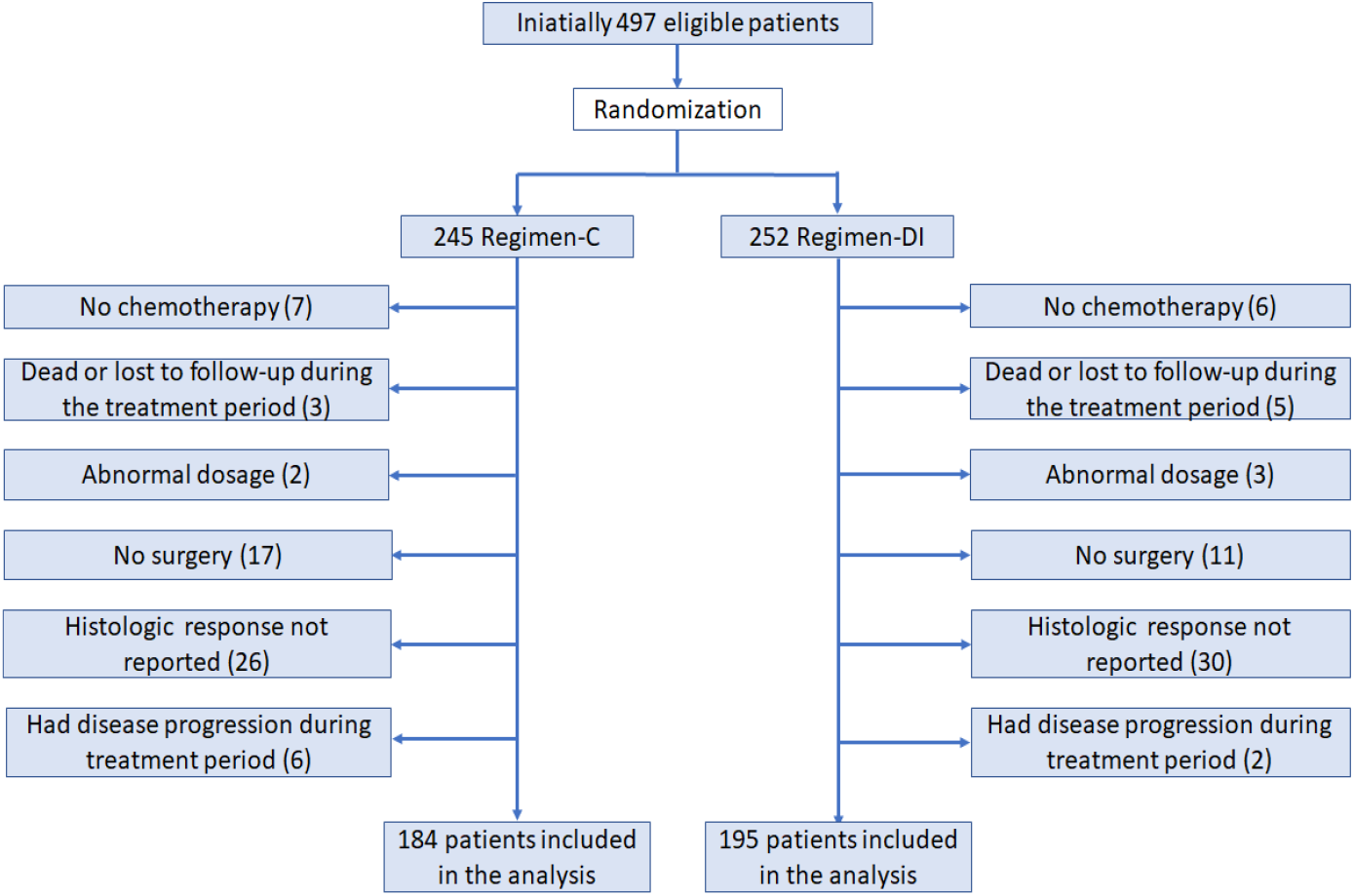
Consort diagram

### Statistical analysis

A mixture cure model^33-35,25^ is used to assess the association of several variables of interest with cure and survival time for the uncured patients. This model assumes that the probability of survival without progression until time *t*, given covariates *x* and *z*, is

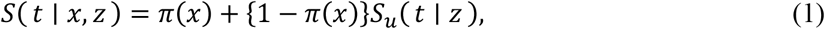

Where *π*(*x*) and *S*_*u*_(*t* | *z*) denote the probability of being cured given the covariate *x* and the probability of survival without progression until time *t* given the covariate *z*, if the patient is not cured. The covariates of the two components in the mixture cure model can be the same or different, allowing for certain prognostic factors to have an affect only on one of these two outcomes. The probability of being cured *π*(*x*) is modeled by logistic regression and the PFS for the uncured patients *S*_*u*_(*t* | *z*) is modeled by a Cox proportional hazards model.^36-38^ A more detailed description about the model is provided in the Supplementary Material. We used the R package smcure^39^ (version 2.0) to compute the estimates of the parameters and their standard errors in the R statistical software environment^40^.

The prognostic values of histologic response, allocated treatment and received dose-intensity are evaluated through univariate and multivariate mixture cure models. The effects of the variables on the cure fraction and on the PFS for the uncured patients are summarized through odds ratios (OR) and hazard ratios (HR) respectively. An OR larger than 1 means higher cure fraction with respect to the reference category, while a HR of less than 1 indicates a reduced risk for the uncured patients (longer PFS). In addition, the Wald-type confidence intervals are computed at the 95% level. For visual inspection, Kaplan-Meier curves from end of treatment for the whole population and stratified according to histologic response and treatment group are also reported.

## Results

Based on the reverse Kaplan-Meier method^41^, median follow-up time was 60 months (quartiles 52-67 months, range 0-116 months) from end of treatment. A total of 105 patients (57%) from Reg-C and 96 (49%) from Reg-DI experienced cancer relapse or death (all deaths were cancer related) during the follow-up period. Only 3 patients (1%) experienced disease progression after 5 years. The largest observed progression time was 5.3 and 7.7 years in Reg-C and Reg-DI respectively. The estimated PFS for all patients (Figure 2.A) reaches a plateau around 7 years or earlier, indicating a consistent fraction of cured patients. There are 178 (47%) censored patients who were reported alive with no progression at the last contact. In total 159 patients (93%) from Reg-C and 172 (88%) from Reg-DI received the complete 6 chemotherapy cycles, while 19 patients (9 and 10 from Reg-C and Reg-DI respectively) did not receive any chemotherapy after surgery. Good histologic response was observed in 68 (37%) and in 97 (50%) Reg-C and Reg-DI patients respectively. The Kaplan-Meier estimator from end of treatment, stratified by histologic response, is shown in Figure 2.B.

**Fig. 2.**
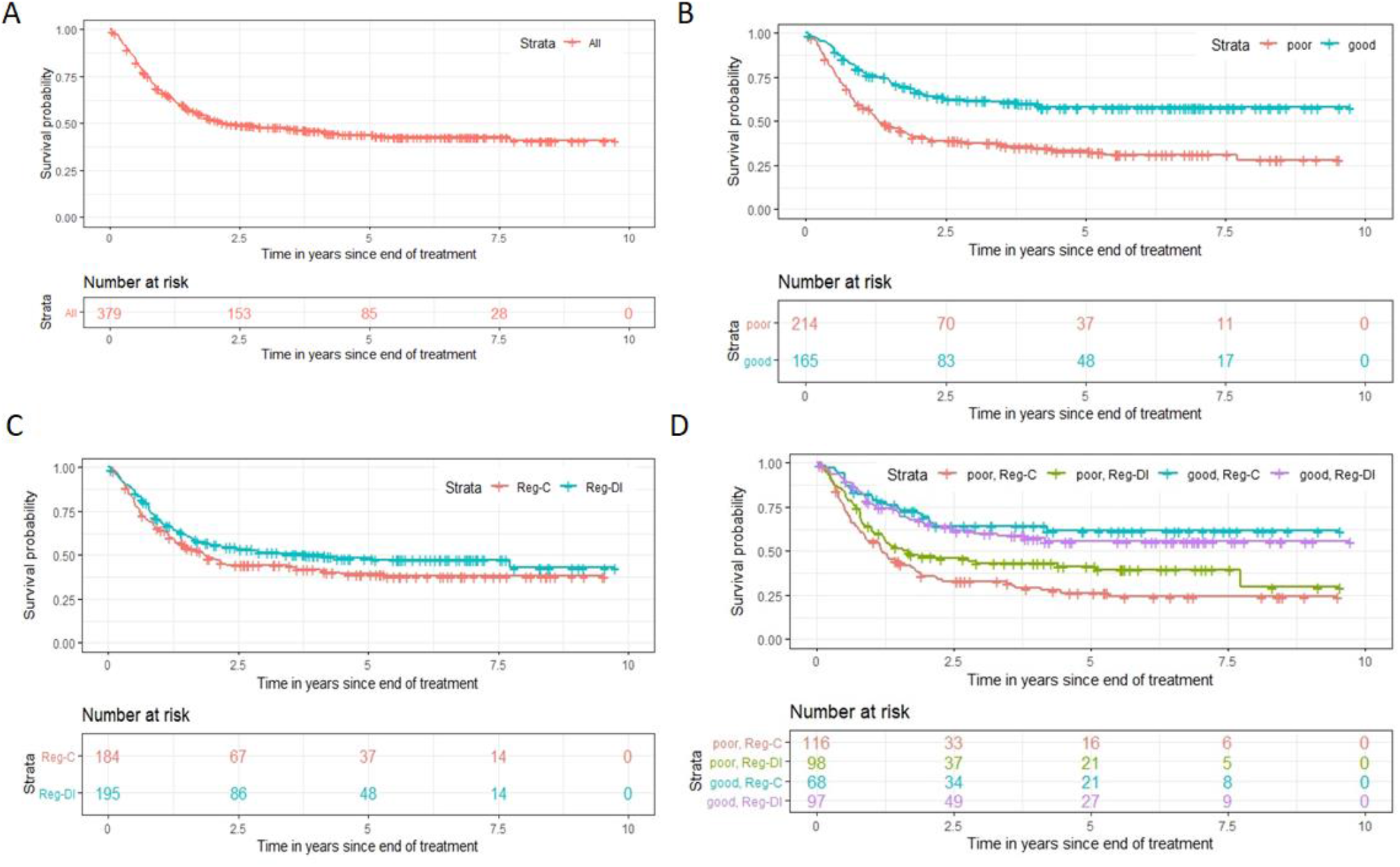
(A) PFS for all the patients. (B) PFS according to histologic response. (C) PFS according to the allocated treatment. (D) PFS according to the allocated treatment and histologic response.

With a univariable logistic/Cox mixture cure model, histologic response is found to be strongly prognostic for the cure fraction (OR: 3.00 [1.75-5.17]) in favor of GR. For the uncured patients, there is no clear indication that good histologic response is associated with longer PFS (HR: 0.78 [0.53-1.16]). The estimated cure fractions and PFS at 3 or 5 years for the uncured patients with good or poor histologic response are shown in Table 1. Apart from separating the long-term from the short-term effect, the mixture cure model provides estimation of PFS for cured and uncured patients combined, as usually done in survival analysis. The estimated 3 and 5-year combined PFS are given in the last two rows of Table 1.

**Table 1.**
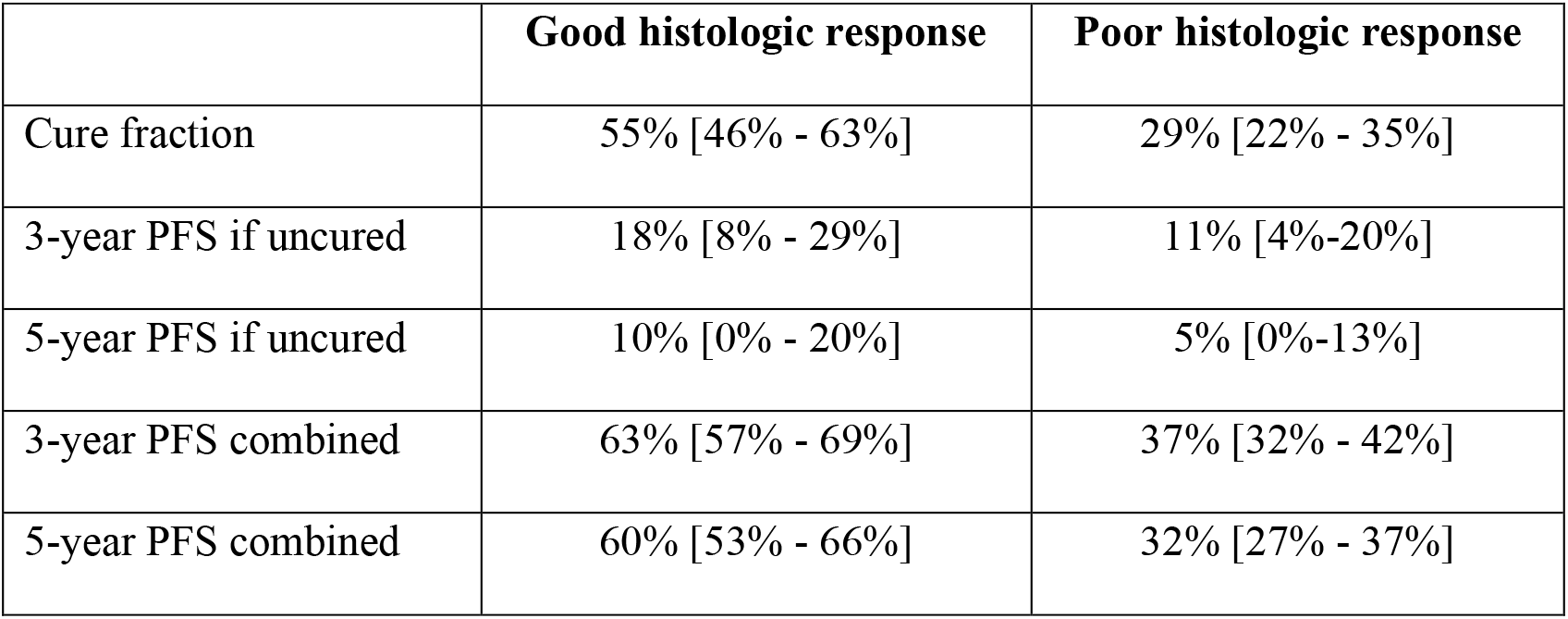
The estimated cure fractions, PFS at 3 and 5 years for the uncured patients, PFS at 3 and 5 years for cured and uncured patients combined according to histologic response. Model based 95% confidence intervals are reported.

A univariable logistic/Cox mixture cure model does not find evidence of a statistically significant effect of the allocated regimen on the cure fraction (OR: 1.35 [0.80-2.29] reference Reg-C). Among the uncured patients, the hazard ratio (HR) is 0.88 [0.59-1.30] in favor of Reg-DI. These estimates suggest that the intensified treatment might be associated with better cure chances and longer PFS, but the confidence intervals are too wide for clear conclusions in both components.

A multivariable cure model is also considered to assess the effect of the treatment group after conditioning on histologic response. A logistic/Cox cure model was fitted with allocated treatment, histologic response and an interaction term as covariates for the cure model. Only histologic response was added to the survival model of the uncured patients to limit the number of parameters that need to be estimated. Good histologic response was found to be strongly associated with good chances of being cured as in the univariable model. Moreover, being allocated to the intensified treatment group (Reg-DI) seems to have a positive effect on the cure fraction among PR (Table 2). Among GR, there is no indication for such a positive effect (Table 2).

**Table 2.** Odds ratios (OR) together with 95% confidence intervals for the effect of the allocated treatment on the cure fraction after conditioning on histologic response.

|  |  | OR [95% CI] |
| --- | --- | --- |
| Poor hist. response | Reg-C |  |
|  | Reg-DI | 1.90 [0.93 – 3.89] |
| Good hist. response | Reg-C |  |
|  | Reg-DI | 0.78 [0.38 – 1.59] |

The estimated cure fractions according to allocated treatment and histologic response are given in Table 3.A. Slightly lower survival for the Reg-DI group versus the Reg-C group among patients with good histologic response can also be observed in the stratified Kaplan-Meier estimator (Figure 2.D). For the uncured patients, good histologic response is associated with longer PFS (HR: 0.78 [0.54-1.13]) compared to the reference group of PR, but not significantly. PFS probabilities at 3 and 5 years for the uncured patients are similar to the univariable model (Table 3s.B). An illustration of the information we obtain with a cure model is provided in Figure 3. Including also the allocated treatment group as a covariate for the survival model of the uncured patients did not show any evidence of an effect of treatment on PFS. A similar behavior is observed if, instead of the allocated treatment, we use the received post-operative dose-intensity defined as in the primary analysis of the trial^8^ (details can be found in the Supplementary Material). This sensitivity analysis was performed since the effect of the pre-operative treatment can already captured in the histologic response.

**Table 3.**
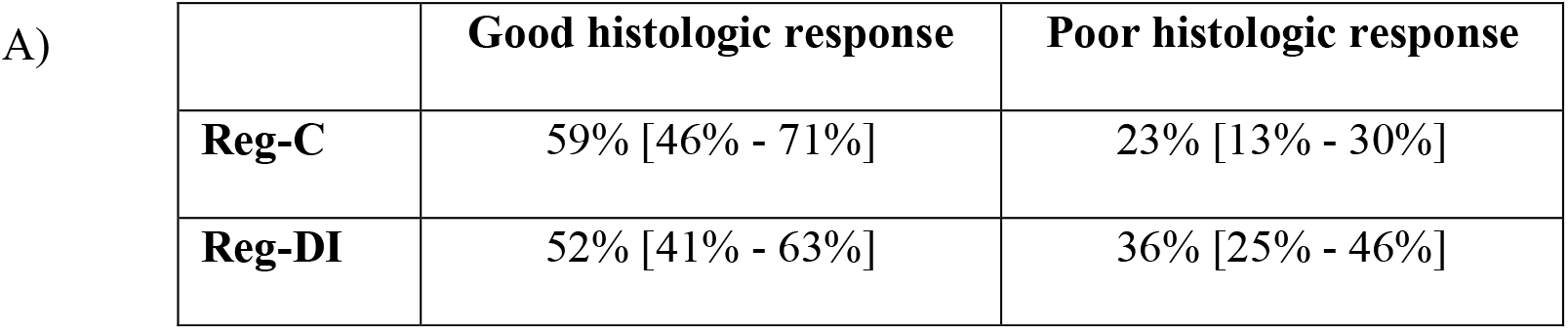
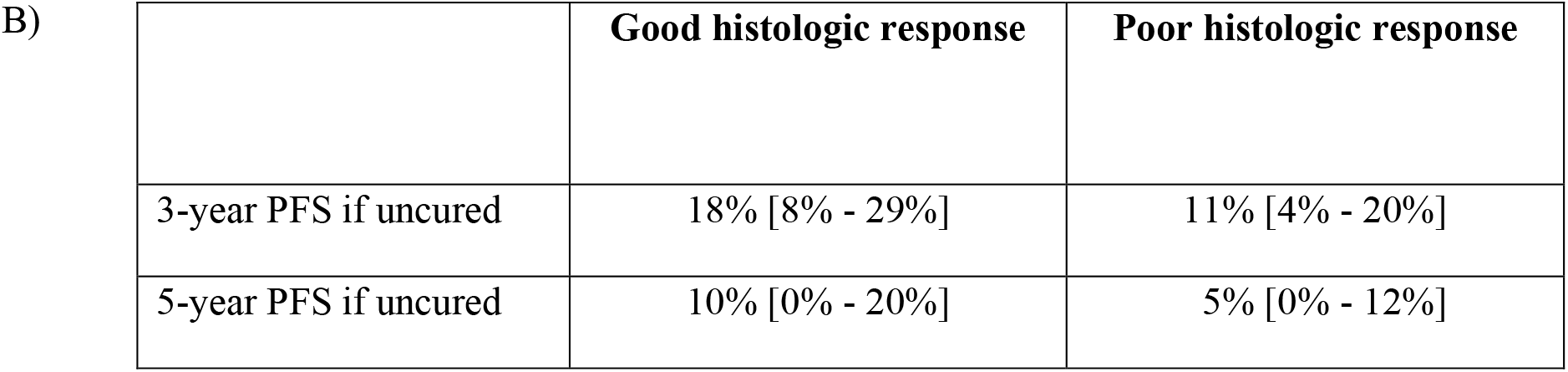
A) Estimated cure fractions and 95% confidence intervals according to allocated treatment and histologic response. B) Estimated 3 and 5-year PFS (and 95% CI) for the uncured patients according to histologic response.

|  | Good histologic response | Poor histologic response |
| --- | --- | --- |
| **Reg-C** | 59% [46% - 71%] | 23% [13% - 30%] |
| **Reg-DI** | 52% [41% - 63%] | 36% [25% - 46%] |
| 3-year PFS if uncured | 18% [8% - 29%] | 11% [4% - 20%] |
| 5-year PFS if uncured | 10% [0% - 20%] | 5% [0% - 12%] |

**Fig. 3.**
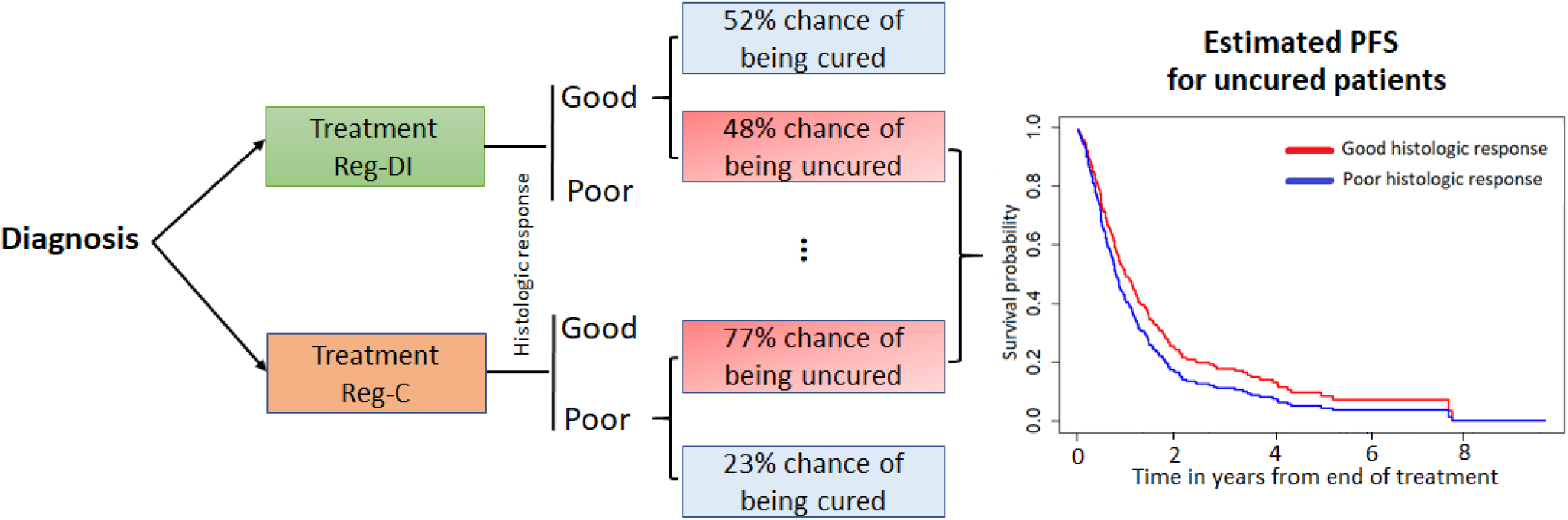
Illustration of cure model interpretation. Cure chances are estimated on the bases of the allocated treatment and the observed histologic response. PFS for the uncured patients is estimated based on histologic response.

## Discussion

The aim of the MRC BO06/EORTC 80931 clinical trial was to investigate whether increasing the dose-intensity would improve the survival of patients with nonmetastatic limb osteosarcoma. The initial analysis of the trial^9^ showed that intensified chemotherapy results in higher chances of good histologic response (GR), mainly related to the increased number of cycles and amount of dose received before surgery. However, even though good histologic response, as prognostic factor, is associated with better survival, no significant better survival was shown in the DI-regimen with a higher proportion of GRs, due to a more intensified treatment. This outcome makes the value of this marker debatable,^11,42-44^ or even better, makes the interpretation of this marker more complex. Here a new point of view is provided on the same data by using a cure model framework that distinguishes between short and long-term effects of covariates.

Previous studies had suggested that a fraction of osteosarcoma patients get cured by treatment, i.e. do not experience cancer progression during their lifetime.^8,10^ However, as result of censoring, it cannot be determined whether patients with relatively short follow-up and last observed alive without showing signs of the disease are cured or not. Compared to the traditional survival analysis methods, which estimate only the survival for the entire cohort, mixture cure models also allow us to estimate the chances of being cured and the PFS if a patient is not cured. From a clinical point of view, the cure fraction might be more informative than 5-year survival rates, in particular for young patients, being the most common patient group with osteosarcoma. Most importantly, enabling the investigation of separate covariate effects on cure and on PFS for the uncured patients, cure models provide a more detailed information of these effects.^21,27,28^

This study showed that the prognostic effect of histologic response on survival is mainly a result of more patients being cured when they have good histologic response. In other words, the chances of being cured after treatment for GR are considerably higher compared to those for patients with poor histologic response (PR), while the effect of histologic response on PFS for the uncured patients was not so clear. With a univariate analysis, there is no evidence of a significant effect of the allocated treatment on cure or on PFS for the uncured patients. However, in a multivariate model accounting for histologic response and allowing for interaction between the treatment arm and histologic response, Reg-DI seemed to have a positive effect on the cure fraction among PR, consistent with the findings of Bishop et al.^10^ The results suggest that, for GR the intensified treatment might have no effect on cure status, i.e. GR in Reg-C do not have lower chances of being cured than GR in Reg-DI, which is also consistent with the findings of Bishop et al.^10^ One explanation might be the following. Since a good histologic response in Reg-C is less common, given that it has been achieved suggests that these responding osteosarcoma cells do not reflect the self-renewing tumorigenic stem cell, at least less than in poor responsive osteosarcoma, hence have a different biologic behavior with higher chances of getting cured.^43^ Patients who respond poorly even despite high DI (Reg-DI) have less chemotherapy susceptible osteosarcoma cells with likely a different molecular profile, resulting in worse cure possibilities. In the initial analysis of the trial^9^, a multivariate Cox regression model, accounting for histologic response, did not find any significant effect for the treatment group or the interaction term. This might be due to a failure to separate short and long-term effect when cured and uncured patients are considered together as one group with the same survival pattern. Not accounting for the presence of cured patients hides some of the treatment effects. Here the highest cure fraction (59%) is observed among GR in Reg-C, while the lowest cure fraction (23%) corresponds to PR in Reg-C.

The focus of this study was on the prognostic value of histologic response and allocated treatment to be in line with the focus of the initial analysis of the trial.^9^ Since the actual received DI differ considerably from the intended ones as result of delays and reductions related to toxicity,^9,14^ the prognostic value of the treatment arm might not properly represent the prognostic value of the intensified chemotherapy. Nevertheless, replacing the allocated treatment by the post-operative received DI gave similar results. The new insights provided by the mixture cure model are relevant for both the patient and clinical perspective. They can help in identifying patients at a higher risk of progression, who might benefit from an intensified chemotherapy, and in avoiding that patients with good chances of being cured undergo a more toxic and aggressive treatment.

## Data Availability

The data that support the findings of this study are available from MRC BO06/EORTC 80931 collaborators and European Osteosarcoma Intergroup but restrictions apply to the availability of these data, which were used under license for the current study, and so are not publicly available.

## Additional Information

Authors’ contributions: EM and MF conceived and designed the study. EM and MF conducted the analysis, and all authors interpreted the results. EM drafted the first version of the manuscript. All authors helped in revising the manuscript and gave their final approval of the submitted version.

