## Supplementary Material for "A new perspective on the BO06 trial in osteosarcoma: short- and long-term prognostic value of histologic response and intensified chemotherapy"

**Mixture cure model**

The idea behind the mixture cure model is to assume that the population consists of two groups: the cured and the susceptible and to combine two different models for simultaneous estimation of the probability of being cured and of the time-to-event if uncured. Two, possibly different, sets of covariates X and Z are used to model the effects of certain prognostic factors separately on cure and time-to-event. If we denote by B an unobserved variable which indicates the cure status, i.e. B=0 if the patient is cured and B=1 otherwise, then the probability of being cured is

$\pi\left( x \right)\mathbb{=P(}B=0\mid X=x)$

and the survival function of the uncured patients is given by

$$S_{u}\left( t \mid z \right)\mathbb{=P}\left( T>t \mid B=1, Z=z \right).$$

It follows that the survival function of a general patient (where the cure status is unknown) is

$$S\left( t \mid x,z \right)=\pi\left( x \right)+\left\{ 1-\pi\left( x \right) \right\}S_{u}\left( t \mid z \right)$$

and the survival reaches a plateau

$$\lim_{t\to\infty} S\left( t | x,z \right)=\pi\left( x \right)>0.$$

Various models (parametric and non-parametric) can be used for both the components but the most common choice is to assume a logistic model for the cure fraction

$$\pi\left( x \right)=\frac{e^{\gamma'x}}{1+e^{\gamma^{'}x}}$$

and a Cox proportional hazards model for the time-to-event of the uncured patients

$$S_{u}\left( t \mid z \right)=\exp(-\Lambda_{0}\left( t \right)e^{\beta^{'}z})$$

for some parameters β, γ and a nonparametric cumulative baseline hazard $\Lambda_{0}.$This model seems to be appropriate for our purposes since it is a good balance between simplicity and flexibility. Note that the proportional hazards assumption is only made at the level of the susceptible patients and not for the whole population. As in the usual logistic and Cox models, the effects of the covariates X and Z are captured through the coefficients γ, β and for interpretation purposes we can compute the odds ratio for being cured and the hazard ratio for the risk of uncured patients.

Since the baseline hazard $\Lambda_{0}$ is left unspecified and estimated nonparametrically, for the model to be identifiable we need sufficient follow-up: the Kaplan-Meier estimator reaches a plateau at a time τ smaller than the maximum follow-up. In other words, the chances for the event of interest to happen beyond the duration of the study should be negligible. Otherwise, the cure rate cannot be correctly identified. We do not need to specify the time τ beyond which the patient can be considered cured, but just to have evidence that there are sufficient observed follow-up times larger than τ.

Estimation of the model is carried out through the maximum likelihood principle. Differently from the common Cox model, because of the unobserved cure status B, it is not possible to derive explicit expressions for the estimators but an iterative Expectation-Maximization algorithm is used to estimate simultaneously β, γ and $\Lambda_{0}.$ The standard errors of the estimated parameters and confidence intervals for the survival at 3 and 5 years are obtained via a nonparametric bootstrap procedure. For our analysis we use 1000 bootstrap samples.

**Received dose-intensity**

Here we evaluate the effect of the received dose-intensity on cure and PFS for the uncured patients accounting for histologic response through a multivariate mixture cure model. Since the effect of chemotherapy intensification during the pre-operative phase is already captured in the observed histologic response, to avoid strong dependencies between variables we consider only the received post-operative dose intensity (Post-DI). As in the initial analysis of the trial, the post-operative dose intensity for both agents (DOX and CDDP) is defined using the formula^9^

Post-DI $=$ $\frac{Received dose after surgery/Expected dose after surgery}{Actual duration of post-operative chemotherapy/Expected duration of post-operative chemotherapy}$

where calculation of the expected dose (mg/m^2^) and duration (days) was based on the protocol for Reg-C, i.e. the expected duration of the post-operative chemotherapy is 84 days (4×3-week cycles) and the expected dose after surgery is 700 mg/m^2^ (4×75 mg/m^2^ DOX + 4×100 mg/m^2^ CDDP). The actual duration of post-operative chemotherapy is computed as the period from the start of the first cycle after surgery until the end of the treatment. The received dose after surgery is computed as the sum of the received doses of DOX and CDDP during all the post-operative chemotherapy cycles, standardized by the body surface area.

For this analysis we further exclude 4 patients, for which at least one of the doses during the post-operative chemotherapy was not reported and 19 patients that interrupted the treatment after surgery, i.e. did not receive any post-operative chemotherapy. As a result, we consider in total 356 patients. Histograms of the Post-DI for each treatment arm and according to histologic response are given in Figure S1. To avoid too many parameters in the model, we consider histologic response, the Post-DI and an interaction factor between them as covariates for the cure status while only the histologic response is used as a prognostic factor for the PFS of the uncured patients. However, including the Post-DI also in the model for the survival of the uncured patients does not find its effect significant. Post-DI is centered at 1, which is the expected post-DI for a patient of Reg-C that receives all the doses as planned.

We again observe that good histologic response is highly associated with good chances of being cured when Post-DI=1 (Table S1). An increase by 0.5 units in Post-DI seems to have a positive effect in the cure fraction among PR, while among GR we observe an opposite effect (Table S1). However, the latter two effects are not found statistically significant and a larger sample would be needed to confirm the results. The estimated cure rates for different combinations of histologic response and Post-DI are given in Table S2. For the uncured patients, as in the main paper, good histologic response is associated with longer PFS (HR: 0.79 [0.55 - 1.14]) compared to the reference group of PR.

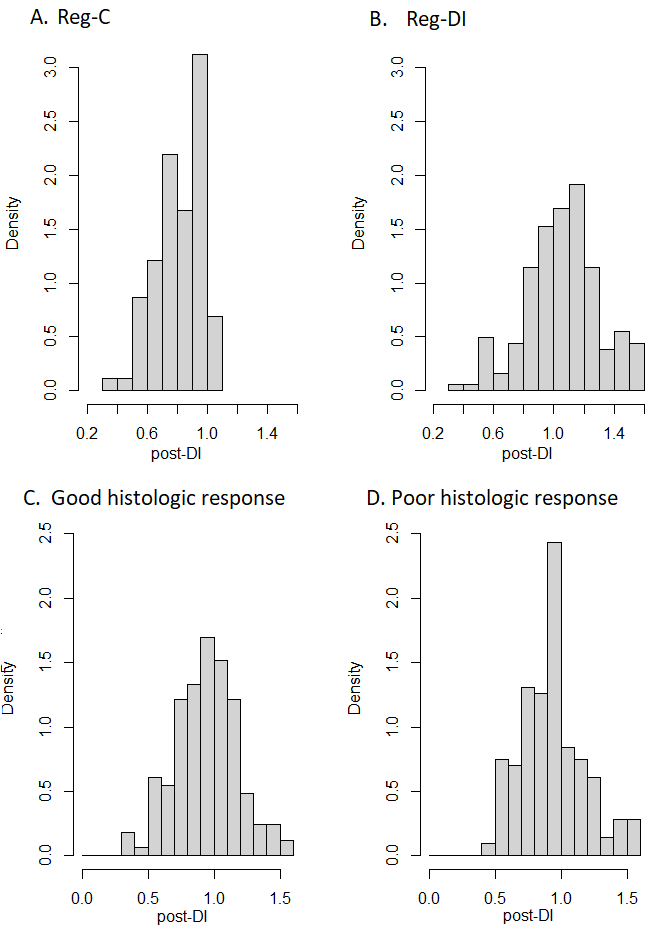

**Fig. S1**. Histogram of the Post-DI according to treatment arm (A and B) and according histologic response (C and D).

|  |  | OR [95% CI] |
| --- | --- | --- |
| Poor histologic response | Post-DI |  |
|  | Post-DI+0.5 | 1.41 [0.61 – 3.25] |
| Good histologic response | Post-DI |  |
|  | Post-DI+0..5 | 0.83 [0.37 – 1.85] |

**Table S1.** The odds ratios (OR) together with 95% confidence intervals for the effect of histologic response and received Post-DI on the cure fraction.

| Post-DI | Good histologic response | Poor histologic response |
| --- | --- | --- |
| 0.5 | 58% [41%, 77%] | 24% [11%, 39%] |
| 1 | 53% [45%, 62%] | 30% [22%, 37%] |
| 1.5 | 48% [28%, 71%] | 38% [17%, 59%] |

**Table S2.** The estimated cure probabilities (and 95% confidence intervals) according to histologic response and Post-DI.
